# PVT in patients presenting with post-acute sequelae of COVID-19

**DOI:** 10.1101/2023.12.14.23299928

**Authors:** Shinjon Chakrabarti, Kamini Krishnan, Rachel Galioto

## Abstract

We investigated performance validity tests (PVTs) in patients presenting with new onset cognitive complaints associated with post-acute sequelae of COVID-19 infection (PASC). Retrospective data were obtained from IRB-approved registries. All patients completed the Victoria Symptom Validity Test (VSVT) in conjunction with a neuropsychological evaluation. A sub-analysis included 7 other PVT measures. The PASC sample was compared to an analogous multiple sclerosis (MS) sample with known PVT failure rates. The PASC sample consisted of 177 patients (49.4 ± 11.2 years), educated (14.7 ± 2.3 years), predominantly female (81.4%), and white, non-Hispanic (85.3%) patients. Seven percent of the PASC sample scored below the established VSVT hard item cut-off, and of those with invalid VSVT over 50% failed 3 or more additional PVTs. In comparison to a MS sample, the PASC sample reported comparable psychological symptoms, but were significantly less likely to produce invalid VSVT scores and seek disability benefits. This study provides a profile of PVTs in patients presenting with PASC. The general infrequence of invalid responding in this PASC sample (7%) is noteworthy compared to an MS sample and highlights the role of additional factors in non-credible response such as elevated psychological symptoms or pursuit of disability.

## INTRODUCTION

The use of performance validity tests (PVTs) during neuropsychological evaluation strengthens the legitimacy of the diagnoses, prognoses, and subsequent clinical recommendations. Factors influencing performance on PVTs can include readily identifiable external incentives, such as applying for disability, and other factors, such as opposition to the evaluation, unnatural fatigue, pain, and psychological conditions, however this largely depends on the patient sample (Sweet et al., 2021). Thus, neuropsychologists routinely administer, in the medicolegal case of disability, forensic settings, and clinical practice, PVTs as part of a standard neuropsychological battery to accurately interpret cognitive data obtained during a neuropsychological evaluation.

Patients recovering from COVID-19 often report cognitive difficulties that persist beyond initial infection. In the COVID-19 literature, this is defined as new or persisting cognitive symptoms present 30 days after COVID-19 diagnosis and referred to as Post-Acute Sequelae of COVID-19 (PASC) (Thaweethai et al., 2023). A recent review summarized the growing research focused on the neuropsychological outcomes of adult survivors of COVID-19 (May, 2022). A majority of the studies analyzed demonstrated the presence of cognitive deficits even in patients without severe infections, where severe is defined by hospitalization at the acute stage or new onset of severe neurological symptoms such as seizures or strokes (May, 2022). However, these studies did no report PVTs. Additional research on PASC patients has similarly suggested the presence of cognitive impairments(Collantes et al., 2021; García-Sánchez et al., 2022; Hampshire et al., 2021). Under the current knowledge of PASC correlated cognitive impairment, it is necessary to strengthen our understanding of performance validity testing, which is often underreported in current literature.

PASC exhibits manifestations and symptomatology including chronic pain, fatigue, cognitive impairment, and psychological distress that resembles fibromyalgia syndrome (FMS) and chronic fatigue syndrome (CFS) (Haider et al., 2023; Plaut, 2023). PVT performance is well-characterized in these groups and as such PASC patients may have similar outcomes. Prior studies indicate invalid performance on PVTs in FMS and CFS occurs in a minority of patients, but at a relatively higher rate than normal (Bar-On Kalfon et al., 2016; R. Gervais, 2004; Teodoro et al., 2018). Additionally, disability seeking patients with FMS and CFS produce significantly higher rates of PVT failure (R. O. Gervais et al., 2001; Teodoro et al., 2018).

However, other studies found low levels of reduced PVT performance indicating the absence of underperformance on cognitive testing (Busichio et al., 2004; Cockshell & Mathias, 2012). In contrast, typical performance on PVTs in patients with PASC is largely unknown. Whiteside and colleagues reported a 7.5% PVT failure rate in their PASC sample, which is consistent with most clinical samples(Whiteside et al., 2021). A similar association may occur in our PASC sample.

In addition to cognitive deficits, patients recovering from COVID-19 are often at risk for psychiatric sequelae (Zhou et al., 2020). In several patient populations, including those with multiple sclerosis (MS) and mild traumatic brain injury, and psychological disorders, and pending disability applications are associated with invalid performance PVTs (Doss et al., 1999; Galioto et al., 2020; Grote et al., 2000). Additionally, vaccine hesitancy is shown to be associated with lower scores on executive function tasks (Batty et al., 2021). The prevalence of invalid performance in patients with PASC and the potential mitigating factors that lend to failed PVT performance have not been investigated and warrant further investigation.

The goal of this study was to examine PVT performance in patients with PASC investigating differences in demographic variables, neuropsychological testing, and disability and vaccination status in patients based predominantly on VSVT performance. We also included a secondary analysis with multiple PVT measures as is recommended in the literature (McWhirter et al., 2020; Sweet et al., 2021). Furthermore, the PASC sample will be compared to additional patient populations including MS, chosen due to similarities in the lifespan prevalence, high incidence of psychological symptoms, disability status, and comparable literature that allows us to compare the PASC sample to well-established base rates of PVT failure.

## METHODS

### Participants and Procedures

Patients with PASC were referred for a neuropsychological evaluation following physician evaluation in a Covid-19 recovery specialty clinic. The following inclusion criteria was used to form the PASC sample 1) adults aged 18 or above with a PCR confirmed COVID-19 infection with onset of cognitive complaints reported only following infection 2) English as the primary language 3) completed a neuropsychological evaluation including the VSVT. We excluded patients with neurological diagnosis prior to developing COVID-19 (such as epilepsy, stroke, MS, etc.)

Data for this retrospective study were obtained from IRB approved clinical neuropsychological registries for MS and COVID-19 within an academic medical center. Comparison sample included patients with MS who were referred for neuropsychological evaluation. This study was conducted in accordance with the Helsinki declaration.

We identified 310 patients with PCR confirmed COVID-19 between March of 2020 and May of 2022. Ninety patients with a pre-existing neurological diagnosis prior to COVID-19 infection were excluded. An additional 43 were excluded because they were either administered a PVT other than VSVT (n=17) or not administered a PVT (n=26). This was a clinical sample, hence contributing to some variability in PVT use by the neuropsychologists in this practice.

### Clinical Variables

Hospitalization during the acute stage of infection and vaccination status for patients with COVID-19 was determined through direct questioning and electronic medical chart review. The number of days between PCR confirmed COVID-19 infection and neuropsychological testing was calculated.

### Neuropsychological Testing

Memory measures included the Rey Auditory Verbal Learning Test (RAVLT; (Rey, 1964) and the Brief Visuospatial Memory Test-Revised (BVMT-R; (Benedict, 1997)). Attention and executive functioning measures included Digit Span from the Wechsler Adult Intelligence Scale-IV (WAIS-IV; (Wechsler, 2009) and Trail Making Test (Reitan & Wolfson, 1985). Processing speed was measured through Coding subtests from the WAIS-IV (Wechsler, 2009).

Neuropsychological data were converted from raw scores to standardized scores based on published normative data (Scaled scores mean-10, SD=3, WAIS-IV digit span and coding; T-scores (mean=50, SD=10, Trail Making Test and BVMT-R; Standard scores (mean=100, SD=15, RAVLT). Dichotomous variables were created (0=not impaired, 1= impaired) with impairment defined as T-scores ≤35, scaled scores ≤5, and Standard scores ≤80), consistent with cutoff for mild impairment (Heaton et al., 1992).

### Performance Validity

The Victoria Symptom Validity Test (VSVT; (Slick et al., 1997) is a two-alternative forced-choice, free standing PVT. The test consists of 24 “easy” and 24 “hard” target, 5-digit number strings that are classified based on the degree of similarity between the targets and foils. A recent cross-validation study noted a ≤ 16 hard item cut-off to have greater specificity compared to other VSVT associated metrics and this was used to determine the specific cutoff scores on this study (Resch et al., 2021).

The Advanced Clinical Solutions: Word Choice Test (WCT; (Pearson, 2009) is a visuoauditory PVT. WCT contains 50 learning items followed by 50 test items. Immediately following the learning trials, patients are asked to distinguish the target words from a series of 50-word pairs containing the original words and a distractor. Pearson (2009) recommends an invalid cutoff of (<45/50).

The Test of Memory Malingering (TOMM; (Tombaugh, 1996) is a forced choice visual recognition performance validity test comprised of two learning trials and a retention trial following a 10-minute delay. Tombaugh (1996) recommends a ≤45 cutoff on learning trial 2 or the retention trial as an indicator of memory malingering.

The Rey Dot Counting Test (DCT) is a brief-time based performance validity test developed by Andrey Rey and described by Lezak(Lezak, 1995). Patients are instructed to count the number of dots on two subsets of cards: grouped and randomly arranged. Response times are measured, and nonvalid scoring is determined when the time taken to count grouped dots is ≤ to the time taken to count ungrouped dots.

Furthermore, five embedded PVT measures with the following cutoffs were included: RAVLT Recognition (≤ 9) (Pliskin et al., 2021), BVMT recognition discrimination (≤ 4) (Pliskin et al., 2021), WAIS-IV reliable digits (≤ 7) (Schroeder et al., 2012), Trail making test A and B (T ≤ 33) (Abeare et al., 2019).

### Psychological Variables

Depression and anxiety were assessed using the Beck Depression Inventory – Second Edition (BDI-II; (Beck et al., 1996) and the Beck Anxiety Inventory (BAI; (Beck et al., 1988). Total scores on both measures range from 0 to 63, and higher scores represent greater symptom severity.

### Disability status

Disability status was assessed for patients with PASC and MS via direct questioning during the evaluation and confirmed by chart review. Disability status was broken down into three groups: (1) patient has not previously applied, is not considering applying, and/or is retired; (2) patient is in the process of applying, has an application under review, or in the process of appealing a recent rejection; or (3) patient is currently on disability (social security disability or long-term disability).

### Statistical Analyses

Descriptive statistics, including demographics and clinical variables, for the PASC sample and comparison to the patients with MS, when applicable, are included in Table 1. Chi-square and independent sample t-tests were used to compare patients who were and were not administered a PVT. Spearman correlations were used to determine the association between VSVT hard item scores and neuropsychological test performance, clinical and demographic variables. Descriptive statistics were used to demonstrate performance on other PVT’s including embedded measures of performance validity. A Mann-Whitney U test and chi-square analyses examined differences in VSVT performance (valid/invalid) on demographic, clinical, and cognitive measures as well as group differences between MS and PASC. Bonferroni correction was applied to two tests with post hoc analyses. A secondary analysis included stand-alone PVT’s and five embedded PVT’s.

**Table 1:**
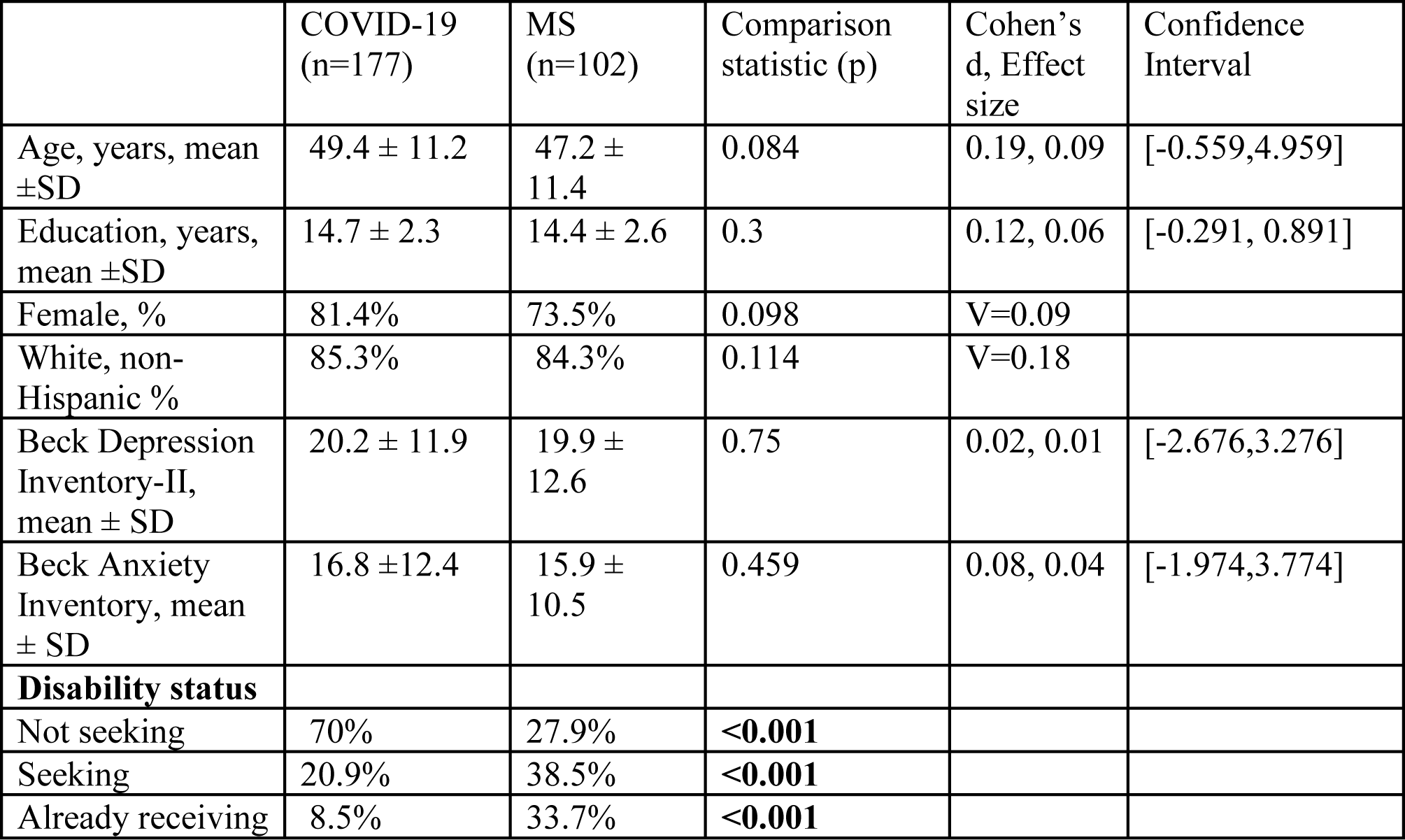
Characteristics of the sample and differences between the groups.

## RESULTS

### Descriptive data for patients with PASC

Independent sample t-tests revealed that patients who were administered a PVT (n=164) were younger, *t* (218) = 8.18, p<0.001, and less educated *t*(218)= 1.829, p=0.034 compared to those who were not administered any PVT. Chi-square analyses revealed a significant difference in disability status between patients who were administered a PVT and those who were not (χ²=6.44, p=0.04). Patients who were not administered a PVT were either working (92%) or disabled (8%).

Table 1 provides sample characteristics. Patients with PASC were middle-aged (49.4 ± 11.2 years), highly educated (14.7 ± 2.3 years), predominantly female (81.4%) and white, non-Hispanic (85.3%). Twenty-four percent of patients required hospitalization at the time of the initial infection. Patients underwent neuropsychological evaluations an average of 8 months after the initial infection. Most patients were not seeking disability (70%). About 8.5% were already receiving disability benefits and 20.9% had a pending disability application at the time of the neuropsychological evaluation (Table 1).

### VSVT performance in patients with PASC

Scores on VSVT ranged from 10 to 24 on the easy items and 10 to 24 on the hard items for the entire sample (n=177). Thirteen patients (7.3%) scored below the VSVT hard item cutoff. Most of those patients (12/13) were administered a second PVT. One hundred percent of those who were administered Word Choice (n=8) performed below the recommended cut-off. Twenty-five percent passed either the TOMM or Dot counting. One patient with 3 PVT’s performed below the recommended cut-off for VSVT and Word Choice and above the cut-off for TOMM.

### Additional PVT’s in patients with PASC

Table 2 provides performance on stand-alone and embedded PVT’s among all patients. Patients with valid VSVT also demonstrated valid performance on another stand-alone PVT such as Word Choice or TOMM (n=118) and 100% demonstrated 3 or fewer failures on embedded measures (Table 3). In the group with invalid VSVT, 12/13 patients failed another stand-alone PVT while 76% of the invalid VSVT sample performed below established cut-offs on 2 or more measures (Table 3). One patient in the invalid VSVT group was not administered any other tests.

**Table 2:** Performance on other performance validity indicators.

|  | Invalid based on VSVT (n=13) | Valid based on VSVT (n=164) |
| --- | --- | --- |
| TOMM<br>Valid<br>Invalid | 2/3<br>1/3 | 3/3<br>0/3 |
| Word Choice<br>Valid<br>Invalid | 4/8<br>4/8 | 115/115<br>0/115 |
| Reliable Digit Span<br>Valid<br>Invalid | 6/12<br>6/12 | 129/163<br>34/163 |
| Trail Making Test A<br>Valid<br>Invalid | 6/12<br>6/12 | 146/159<br>13/159 |
| Trail Making Test B<br>Valid<br>Invalid | 7/12<br>5/12 | 145/159<br>14/159 |
| Rey AVLT recognition<br>Valid<br>Invalid | 4/8<br>4/8 | 152/155<br>3/155 |
| BVMT-R RD<br>Valid<br>Invalid | 3/8<br>5/8 | 139/152<br>13/152 |

Note: Two patients in the invalid group were administered the Dot Counting test and performed below expectations. This test was not given to anyone in the Valid VSVT group and is not reported in the table above.

**Table 3:**
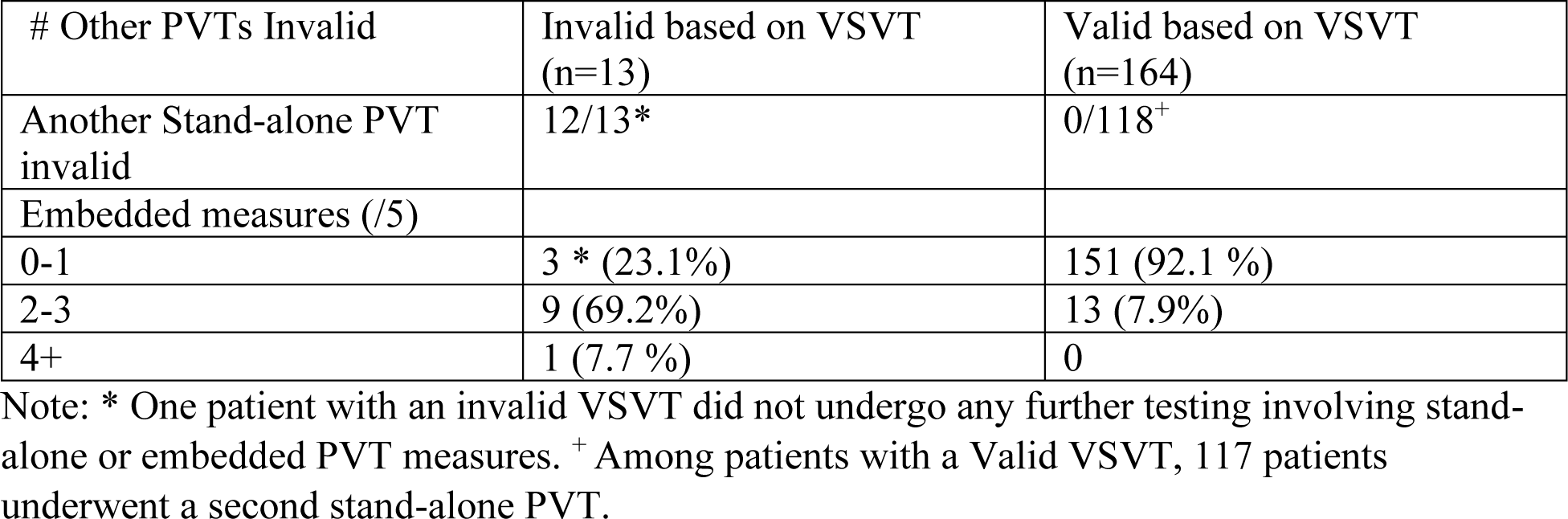
Failure count on stand-alone PVT’s and embedded PVT’s based on VSVT hard item cut-off.

### Association between VSVT and characteristics of patients with PASC

VSVT hard item score was significantly correlated with education (r = 0.17, p = 0.027). Spearman correlation analyses showed significant associations between VSVT hard item raw score and performance on all neuropsychological tests except BVMT (p’s range from 0.050 to <0.001; see Table 4. Spearman correlation coefficients were significant between VSVT hard item raw score and BDI-II (r = -.031, p <0.001), and BAI (r = -0.29, p <0.001). There were no significant associations with other demographic or clinical variables (See Table 4).

**Table 4:** Spearman correlation coefficients between VSVT hard item raw score and demographic, clinical, and cognitive variables.

| Variable | R | P |
| --- | --- | --- |
| Age | 0.04 | 0.60 |
| Education | 0.17 | **0.027** |
| Female | 0.12 | 0.11 |
| White | -0.02 | 0.80 |
| BDI-II | -0.31 | **<0.001** |
| BAI | -0.29 | **<0.001** |
| Time between COVID-19 infection and Neuropsych evaluation | -0.13 | 0.86 |
| Hospitalized | -0.05 | 0.49 |
| COVID Vaccination | -0.04 | 0.65 |
| Disability status | -0.36 | **<0.001** |
| Neuropsychological tests |  |  |
| WAIS-IV: Digit Span | 0.37 | **<0.001** |
| WAIS-IV: Coding | 0.29 | **<0.001** |
| Trail Making Test A | 0.29 | **<0.001** |
| Trail Making Test B | 0.24 | **0.002** |
| RAVLT total | 0.15 | **0.050** |
| RAVLT delay | 0.19 | **0.015** |
| BVMT-R Total | 0.098 | 0.22 |
| BVMT-R delay | 0.13 | 0.10 |

Note: BDI-II: Beck Depression Inventory- Second Edition; BAI: Beck Anxiety Inventory; WAIS-IV: Wechsler Adult Intelligence Scale- Fourth Edition; RAVLT: Rey Auditory Verbal Learning Test; BVMT-R: Brief Visuospatial Memory Test-Revised.

### Comparison of Valid and Invalid performance on the VSVT

#### Neuropsychological test performance

Given the association between VSVT, demographic, clinical, and cognitive measures, we evaluated differences between valid and invalid response on VSVT hard item scores as well as differences in rates of cognitive impairment in these groups. As expected, patients with invalid VSVT scores performed significantly lower across all neuropsychological tests administered when compared to patients with valid scores, except for RAVLT delayed recall (see Table 5). Patients with invalid VSVT demonstrated lower rates of impairment across all cognitive measures and reported greater psychiatric symptoms on BDI-II (U=491, dCohen= 0.501, *η^2^=0.059*) and BAI (U=668, dCohen= 0.341, *η^2^=0.28*). There were no significant differences in demographic variables, rates of hospitalization, or time between PCR-confirmed infection and neuropsychological evaluation between the PASC patients with valid or invalid VSVT hard item scores (See Table 5).

**Table 5:** Characteristics by VSVT outcome in patients with COVID-19.

| Variable | Valid | Invalid | p | U | $d_{\text{Cohen}}, \eta^2$ | Confidence interval |
| --- | --- | --- | --- | --- | --- | --- |
| Age | 49.64 ± 11.14 | 47.08 ± 11.76 | 0.353 | 1231 | 0.14,0.005 | [-10,4] |
| Education | 14.72 ± 2.27 | 14.31 ± 2.02 | 0.55 | 1170 | 0.088,0.002 | [-2,1] |
| Female | 80% | 92% | 0.29 |  | V=0.79 |  |
| White | 85% | 85% | 0.20 |  | V=0.18 |  |
| BDI-II | 19.26 ± 11.40 | 31.69 ± 12.67 | **<0.001** | 491 | 0.501,0.059 | [1,18] |
| BAI | 16.13 ± 11.95 | 25.23 ± 15.01 | **0.029** | 668 | .341,0.28 | [5,21] |
| Time between COVID-19 infection and Neuropsych evaluation | 247.85 ± 141.32 | 243.77 ± 148.44 | 0.92 | 1100 | 0.29,0 | [-83,73] |
| Hospitalized (% yes) | 24% | 23% | 0.92 |  | V=0.008 |  |
| COVID-19 Vaccination (% not vaccinated before infection) | 75% | 53.9% | **0.049** |  | **V=0.24** |  |
| % declined vaccination | 13.4% | 46.1% | **<0.001** |  |  |  |
| Disability status (% pending disability) | 17% | 69% | **<0.001** |  | **V=0.36** |  |
| **Neuropsychological tests, mean ± SD** |  |  |  |  |  |  |
| WAIS-IV Digit Span (n=163/12) Scaled score | 9.72± 2.71 | 6.0 ± 3.1 | **<0.001** | **1591** | **0.569,0.075** | **[-5,-2]** |
| WAIS-IV Coding (n=154/6) Scaled score | 10.05± 2.4 | 6.5± 3.08 | **0.009** | **751** | **0.419,0.042** | **[-6,-1]** |
| Trail Making Test A (n=159/12) T-score | 49.31± 11.6 | 33.25± 18.49 | **0.005** | **1418** | **0.439,0.046** | **[-26,-5]** |
| Trail Making Test B (n=159/12) T-score | 48.89 ±11.52 | 31.33± 18.97 | **<0.001** | **1521** | **0.543,0.069** | **[-27,-7]** |
| RAVLT total (n=157/8) Standard Score | 99.75±1 5.54 | 86.06± 26.89 | 0.085 | 855 | 1.242,0.278 | [-31.76,3.20] |
| RAVLT delay (n=157/8) Standard Score | 103.67± 56.53 | 81.88± 25.98 | 0.72 | 860 | 0.284,0.02 | [-34.98,1.83] |
| BVMT-R Total<br>(n=151/8) T-score | 48.79 ±<br>11.89 | 26.75<br>±9.33 | **0.006** | **959.5** | **0.446,0.047** | **[-20,-4]** |
| BVMT-R delay<br>(n=151/8) T-score | 50.36 ±<br>12.37 | 38.88 ±<br>12.72 | **0.016** | **908.5** | **0.388,0.036** | **[-23,-3]** |
| **Neuropsycholog**<br>**y rates of**<br>**impairment** |  |  |  |  |  |  |
| WAIS-IV Digit<br>Span (n=163/12) | 3.6% | 50% | **<0.001** |  |  |  |
| WAIS-IV<br>Coding<br>(n=154/6) | 1.2% | 42.8% | **<0.001** |  |  |  |
| Trail Making<br>Test A<br>(n=159/12) | 12.2% | 46% | **0.002** |  |  |  |
| Trail Making<br>Test B<br>(n=159/12) | 12.2% | 46% | **0.002** |  |  |  |
| RAVLT total<br>(n=157/8) | 9.2% | 30.8% | **<0.001** |  |  |  |
| RAVLT delay<br>(n=157/8) | 11.6% | 30.8% | **<0.001** |  |  |  |
| BVMT-R Total<br>(n=151/8) | 14.6% | 23.1% | **<0.001** |  |  |  |
| BVMT-R delay<br>(n=151/8) | 12.8% | 30.8% | **<0.001** |  |  |  |

Note: BDI-II: Beck Depression Inventory- Second Edition; BAI: Beck Anxiety Inventory; WAIS-IV: Wechsler Adult Intelligence Scale- Fourth Edition; RAVLT: Rey Auditory Verbal Learning Test; BVMT-R: Brief Visuospatial Memory Test-Revised.

#### Disability status

A Kruskal-Wallis H test showed a statistically significant difference in VSVT hard item raw score by disability status, χ2(2) = 38.48, p < 0.001, with a mean rank score of 103.82 for patients working, 53.64 for those pending disability and 52.77 for patients already on disability. Post hoc analysis showed patients with PASC applying for disability scored significantly lower on VSVT hard items (M = 18, SD = 6.48) compared to those who were not applying (M = 22.7, SD = 2.3; p<0.0001 after Bonferroni correction, Cohen’s d = 0.54). Furthermore, patients already receiving disability benefits scored significantly lower on VSVT hard item score (M=21.49, SD = 4.1) compared to those not applying (M=22.7, SD = 2.3, p= 0.043 after Bonferroni correction; Cohen’s d=0.53). Patients with invalid scores were more likely to have a pending disability application compared to patients with valid VSVT scores (p < 0.001). (See Table 6).

**Table 6:** VSVT hard item cut-off in COVID-19 and MS groups.

|  | COVID-19 (n=177)<br>Count (%) | MS (n=102)<br>Count (%) |  |
| --- | --- | --- | --- |
|  | 13 | 15 |  |
| Not seeking | 2/126 (1.5%) | 2/29 (6.9%) | **<0.001** |
| seeking | 10/37 (27%) | 9/39 (22.5%) | 0.20 |
| Already receiving | 1/15 (6%) | 4/34 (11.4%) | 0.055 |

#### Vaccination Status

Patients were divided into the following groups 1) COVID-19 infection before first dose of the vaccine (n=130) 2) COVID-19 infection after full vaccination (n=10) 3) patient refused COVID- 19 vaccine (n=28) 4) no information available regarding vaccination status (n=9). There were significant differences in patients with valid VSVT performance by vaccination status (χ2=27.28, p<0.001). Patients who declined COVID-19 vaccination scored significantly lower (M= 19.50, SD= 5.9) than patients who developed COVID-19 prior to receiving the first dose of the vaccine (M= 21.91, SD= 3.73, p= 0.03 after Bonferroni correction). Patients with invalid VSVT scores declined vaccination or were unvaccinated at higher rates compared to patients with valid performance (Table 5).

#### Comparison to MS sample

One-way ANOVA revealed no significant differences between MS and PASC in terms of demographic and mood symptoms. Patients with MS produced invalid VSVT scores at a significantly higher rate (14.7%) than patients with PASC (7. 3%), while patients with PASC were less likely to seek disability (Table 6).

## DISCUSSION

This is the first paper to specifically examine PVT performance among patients presenting with cognitive complaints following a PCR-confirmed COVID-19 infection, also known as PASC. We found relatively low rates of invalid performance (7.3%) and uncovered some interesting trends about 1) differences with MS patients 2) disability, 3) vaccination status and 4) standard PVT use. Additionally, performance on the VSVT was not associated with COVID-19 disease markers (hospitalization, duration of symptoms).

First, as expected, there were no differences in demographic variables between the PASC and MS sample. However, it is interesting that both groups report comparable levels of depression and anxiety symptoms, further highlighting the similarities in pronounced levels of psychological symptom endorsement, a known risk factor for invalid performance (Galioto et al., 2020). In this study, seven percent of patients with PASC failed PVT while 14% of patients with MS demonstrated invalid performance, which suggests that the PASC sample was significantly less likely to seek disability benefits compared to the MS sample. Currently, the cognitive manifestations of MS are better understood than COVID-19. However, the largely valid PVT performance in the PASC group provides stronger validation to the common cognitive difficulties reported by recovering patients. Additionally, the results of the PASC sample contrast with findings in MS literature, where patients applying for disability were 6.75 times more likely to produce an invalid score on the administered PVT (VSVT) compared to those who were not applying for disability (Galioto et al., 2020). Furthermore, the lower rate of disability seeking may be accounted for by the low suspicion of prolonged cognitive deficits in most patients with PASC, compared to the consistent neurological, physical, and cognitive symptoms associated with the diagnosis of MS. It should be noted that the PASC subgroup with invalid VSVT performance endorsed higher levels of depression and anxiety compared to patients with valid performance, which may suggest symptom exaggeration or high psychological distress impacting cognition.

The rate of PVT failure in this PASC sample is lower than published rates for fibromyalgia and CFS. Bar-On Kalfon (2016) reported 16% failure in patients with FM and Suhr (2003) comparably found 18.5% and 17.9% failure in CSF and FM patients respectively, which is higher than the current study’s seven percent failure on VSVT. Additionally, psychological symptom endorsement was not significantly elevated in FM patients with low levels of effort, which contrasts the significant difference found in the PASC sample (Bar-On Kalfon et al., 2016). However, disability seeking was similarly associated with PVT failure in all patient groups. Gervais (2001) reported incomplete effort in 35% of FM patients who were on or seeking disability, which is comparable to the 20.9% found in the PASC sample.

Second, it is striking that 20.9% of patients are seeking disability specifically related to sequelae from COVID-19. In a sample restricted to patients with new onset cognitive complaints post infection suggests that a subset of patients with PCR confirmed Covid-19 are experiencing significant difficulties, including cognitive deficits, which have altered their occupational trajectory. This finding is consistent with recent papers that demonstrate chronic cognitive deficits in patients regardless of the severity of COVID-19 infection (García-Sánchez et al., 2022; Krishnan et al., 2022). Further investigations to determine whether cognitive, mood, and/or physical sequelae of COVID-19 contribute to disability are warranted as it can have far-reaching implications for the economic and financial cost of COVID-19 to the patient and society.

Third, it is very interesting, and unexpected, that declining the COVID-19 vaccine was strongly associated with lower VSVT hard item scores and a greater likelihood of non-credible performance. The reasons underlying this association remain unclear, but relatively high rates of pending disability applications (40%) and/or greater severity of initial illness (25% requiring hospitalization) among patients who declined vaccination may be contributory. Studies have demonstrated greater vaccine hesitancy in individuals with poor executive function or lower cognitive function (Acar-Burkay & Cristian, 2022; Batty et al., 2021). The association between PVT failure and lower premorbid intelligence quotient (IQ) has been demonstrated recently (Soble et al., 2023). However, it is difficult to investigate the association between premorbid IQ and vaccine hesitancy in the invalid VSVT group as not all patients underwent IQ testing. Future studies are warranted to investigate this association.

Lastly, consistent with other literature, this study highlights the importance of utilizing multiple PVT’s to inform interpretation (McWhirter et al., 2020; Sherman et al., 2020; Sweet et al., 2021). In this sample, there was a 100% concordance between invalid scores on VSVT and Word Choice. Seventy-seven percent of the PASC sample with invalid VSVT had 2 or more failures on embedded measures of performance validity which was significantly lower than the eight percent failure in the valid VSVT sample.

This study was subject to limitations. Generalization of these results is restricted by the small number of patients with non-credible performance on the VSVT in both the overall sample and each disability subcategory. Furthermore, seeking disability appeared to be a strong determinant of VSVT failure, however with only 21% of our sample seeking disability, this may be an overestimation. Our analyses were also subject to variability in which PVT was administered as this was a clinical sample without consistent test batteries and a priori decisions on managing cases with multiple PVT failures. This sample reflects patients seen during the active pandemic, both pre and immediately post availability of vaccinations. The PVT failure rate may not generalize to patients seen after May 2022.

In conclusion, this study demonstrates that non-credible performance on PVT when using the VSVT, are relatively uncommon among patients with cognitive complaints post-COVID-19. Similar to findings in MS and mild traumatic brain injury literature, psychological factors and disability status are associated with PVT failure. The present study recommends the use of PVTs to disbar the possibility of invalid performance when evaluating cognition in patients with PASC. While invalid performance occurs in a sizeable minority of patients in this sample, we recommend investigating psychological symptoms, disability and vaccination status when conducting neuropsychological evaluations in patients with cognitive complaints post-COVID- 19.

## ACKNOWLEDGMENT

None.

## FUDNING

This work received no funding.

## DATA AVAILABILITY

The data that support the findings of this study are available from the corresponding author, KK, upon reasonable request.

